## Supplementary figures and images for "The Zipime-Weka-Schista study protocol: a longitudinal cohort study and economic evaluation of an integrated home-based approach for genital multi-pathogen screening in women, including female genital schistosomiasis, HPV, Trichomonas and HIV in Zambia"

### 3D model for self-sampling

#### Slide 1
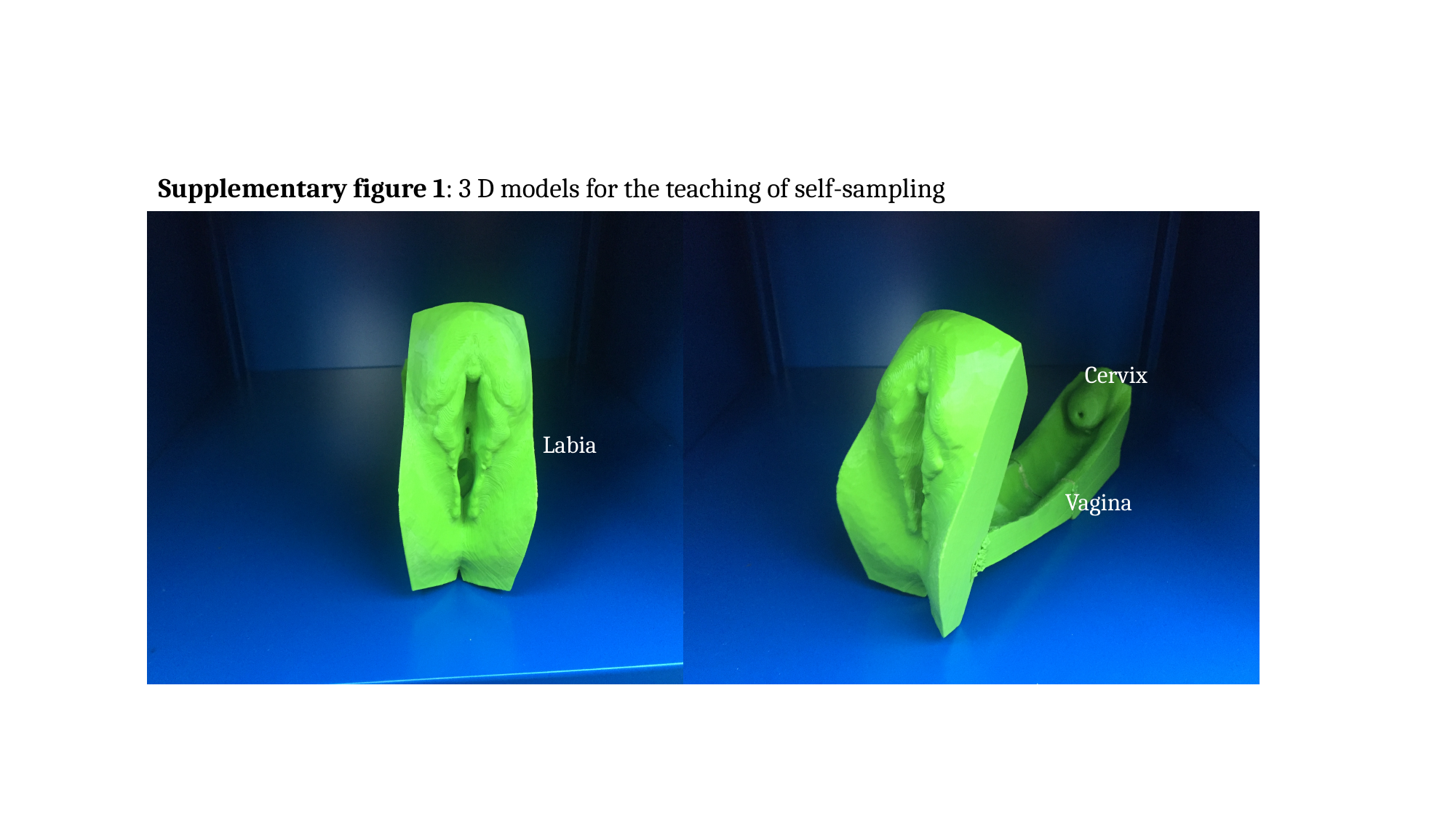

Supplementary figure 1: 3 D models for the teaching of self-sampling
Labia
Cervix
Vagina
